# Analysis of the Number of Tests, the Positivity Rate, and Their Dependency Structure during COVID-19 Pandemic

**DOI:** 10.1101/2021.04.20.21255796

**Authors:** Babak Jamshidi, Hakim Bekrizadeh, Shahriar Jamshidi Zargaran, Mansour Rezaei

## Abstract

**Background:** Applying recent advances in medical instruments, information technology, and unprecedented data sharing into COVID-19 research revolutionized medical sciences, and causes some unprecedented analyses, discussions, and models.

**Methods:** Modeling of this dependency is done using four classes of copulas: Clayton, Frank, Gumbel, and FGM. The estimation of the parameters of the copulas is obtained using the maximum likelihood method. To evaluate the goodness of fit of the copulas, we calculate AIC. All computations are conducted on Matlab R2015b, R 4.0.3, Maple 2018a, and EasyFit 5.6, and the plots are created on software Matlab R2015b and R 4.0.3.

**Results:** As time passes, the number of tests increases, and the positivity rate becomes lower. The epidemic peaks are occasions that violate the stated general rule –due to the early growth of the number of tests. If we divide data of each country into peaks and otherwise, about both of them, the rising number of tests is accompanied by decreasing the positivity rate.

**Conclusion:** The positivity rate can be considered a representative of the level of the spreading. Approaching zero positivity rate is a good criterion to scale the success of a health care system in fighting against an epidemic. We expect that if the number of tests is great enough, the positivity rate does not depend on the number of tests. Accordingly, the number and accuracy of tests can play a vital role in the quality level of epidemic data.

**Key messages:**

- In a country, increasing the positivity rate is more representative than increasing the number of tests to warn about an epidemic peak.
- Approaching zero positivity rate is a good criterion to scale the success of a health care system in fighting against an epidemic.
- Except for the first half of the epidemic peaks, in a country, the higher number of tests is associated with a lower positivity rate.
- In countries with high test per million, there is no significant dependency between the number of tests and positivity rate.

## Introduction

Dr. Li Wenliang, a 34-year-old ophthalmologist, warned his colleagues and set the alarm to the society about a new infection caused by a type of coronavirus in December 2019 in Wuhan, China [1]. Shortly after his warning, all over the world encountered this epidemic. WHO declared this fast speeding infection (COVID-19) in March 2020. As of January 27, 2021, over 100 million cases, and around 2200 K deaths involving COVID-19 have been reported around the world.

The epidemic COVID-19 is the most informative pandemic throughout history. These unprecedented recorded data give rise to some unprecedented concepts, relationships, analyses, discussions, and models [2]. Modeling the dependence between the number of tests and the proportion of positivity (positivity rate) is one of these new issues.

The proportion of positivity is a critical measure because it gives us an indication of how widespread infection is in the area of interest. The proportion of positivity helps public health officials answer questions such as:

- What is the current level of SARS-CoV-2 (coronavirus) transmission in the community?
- Are we doing enough testing for the people who are getting infected? [3]

According to the ratio nature, the high proportion of positivity is due to the high number of positive tests or the low number of total tests. Based on the first possibility, a higher positivity rate suggests higher transmission and that there are likely more people with coronavirus in the community who have not been tested yet. On the other hand, according to the second possibility, a high percentage of positivity means that more testing should probably be done. Accordingly, for policymakers, the high value for this parameter suggests either it is not a good time to relax restrictions aimed at reducing transmission, or it is a good time to add restrictions to slow the spread of disease [3]. In this regard, an analytic report segregated by regions in the UK was presented by the Office for National Statistics [4].

This study aims to investigate the time series of positivity rates individually and together with the time series of the number of tests. This investigation is conducted in two analytic methods: regional and temporal. The individual analysis is mainly undertaken based on the peaks of the spreading of the pandemic (Table 3). For the regional aspect, among the 221 countries, we selected twelve countries: the USA, India, the UK, Italy, Iran, the UAE, Bolivia, Guatemala, Nigeria, Australia, South Korea, and South Africa. The reasons for selecting these twelve countries are They are the top countries in the influential indices (Table 1).

**Table 1.** The information on the influential indicators of COVID-19 in the twelve countries of interest.

| Country | Total cases | Total deaths | Total tests | Cases per Million | Deaths per Million | Tests per Million | Population |
| --- | --- | --- | --- | --- | --- | --- | --- |
| USA | 26 M (1) | 429 K (1) | 299 M (1) | 77 K (7) | 1293 (11) | 901 K (19) | 333 M (3) |
| India | 11 M (2) | 154 K (3) | 192 M (2) | **7688 (115)** | 111 (104) | 138 K (107) | 1387 M (2) |
| UK | 3647 K (5) | 98 K (5) | 67 M (5) | 54 K (25) | 1438 (5) | 987 K (17) | 68 M (21) |
| Italy | 2467 K (8) | 85 K (6) | 31 M (8) | 41 K (36) | 1415 (7) | 512 K (40) | 60 M (24) |
| Iran | 1373 K (16) | 57 K (9) | 8635 K (20) | 16 K (88) | 678 (43) | **102 K (126)** | 85 M (18) |
| UAE | 278 K (43) | 792 (90) | 25 M (12) | 28 K (63) | 80 (117) | 2466 K (5) | 10 M (92) |
| Bolivia | 200 K (53) | 9923 (33) | 514 K (106) | 17 K (84) | 844 (34) | **44 K (150)** | 12 M (79) |
| Guatemala | 154 K (67) | 5465 (43) | 738 K (94) | 8519 (112) | 302 (73) | **41 K (153)** | 18 M (65) |
| Nigeria | 122 K (76) | 1504 (80) | 1241 K (75) | **582 (180)** | **7 (178)** | **5939 (194)** | 209 M (7) |
| Australia | 29 K (107) | 909 (88) | 13 M (14) | **1121 (163)** | **35 (138)** | 495 K (41) | 26 M (54) |
| S Korea | 75 K (86) | 1360 (82) | 5284 K (37) | **1500 (155)** | **27 (148)** | **108 K (121)** | 51 M (28) |
| S Africa | 1418 K (15) | 41 K (14) | 7993 K (22) | 24 K (75) | 703 (42) | 137 K (109) | 60 M (25) |

The numbers in parentheses indicate the rank of countries among 221 countries in the world. For example, Iran has the 18<sup>th</sup> population among all countries.
The bold numbers display the ranks of the second half of the global ranking. The ranks 111 to 221 are considered the second half. For example, regarding cases per million, India is a country of the second half.
The light-highlighted cells show that the country is among the highest quarter of the countries based on the relevant parameter. The darker highlighted cells indicate the country is among the top 5% worldwide. For example, the first row illustrates that except for the criterion of the number of tests per million –where it is among the highest quartile–, the USA is of the top 5% in all indicators.

- Some of them are widely different from the others in some indices (Table 1).
- Their positivity rates are greatly dispersed (Table 2).
- The numbers and time of peaks are different about them (Table 3).
- Their quality of health care systems are of different levels.
- Their data, especially about the number of tests are relatively well recorded.
- They are selected from all continents: the USA and Guatemala from North America, Bolivia from South America, India, Iran, the UAE, and South Korea from Asia, the UK and Italy from Europe, Nigeria and South Africa from Africa, and Australia from Australia.

**Table 2.** The properties of the datasets of the countries of interest.

| Country | Lag | Positivity rate | Start | End | Number of days |
| --- | --- | --- | --- | --- | --- |
| USA | 2 | 8 | 27 February 2020 | 27 January 2021 | 334 |
| India | 3 | 6 | 3 March 2020 | 27 January 2021 | 329 |
| UK | 2 | 5 | 21 February 2020 | 25 January 2021 | 338 |
| Italy | 3 | 8 | 19 February 2020 | 27 January 2021 | 339 |
| Iran | 4 | 16 | 1 April 2020 | 26 January 2021 | 303 |
| UAE | 3 | 1 | 11 March 2020 | 26 January 2021 | 318 |
| Bolivia | 5 | 39 | 19 March 2020 | 24 January 2021 | 308 |
| Guatemala | 5 | 21 | 14 March 2020 | 27 January 2021 | 316 |
| Nigeria | 5 | 10 | 17 March 2020 | 27 January 2021 | 313 |
| Australia | 2 | 0.02 | 22 March 2020 | 26 January 2021 | 308 |
| S Korea | 2 | 0.01 | 18 February 2020 | 30 January 2021 | 347 |
| S Africa | 4 | 2 | 14 March 2020 | 30 January 2021 | 320 |

Finally, to illustrate the dependency of the number of tests and the positivity rates, we apply copulas.

Sklar introduced the concept of copulas in 1959 [5]. A copula –mainly parametric, partially semi-parametric, and rarely non-parametric-is a function that completely describes the dependency structure. It contains all the information to link the marginal distributions to their joint distribution. Accordingly, to obtain a valid multivariate distribution function, it suffices to combine several marginal distribution functions with any candidate for the copula function. Thus, for the purposes of statistical modeling, it is desirable to have a large collection of copulas at one’s disposal. Copula is widely applied in diverse fields, including environmental studies [6 –7] finance [8 –9], hydrology [10], and medical studies [11 –16].

### Data

The main data sources of the paper are the website Worldometers [17] and Our World in Data [18]. We summarize and illustrate all the relevant information about the twelve countries in three (twelve-row) tables and three (twelve-partitioned) figures created on Matlab R2015b.

Table 1 includes the key general indicators up to January 25, 2021. It is worth saying that the total indicators or even per-million indicators do not determine the quality of health care systems because there are observable underreported statistics about the countries Bolivia, Guatemala, Nigeria, Iran, and even India. Despite the mentioned reality, we consider the indicator of the number of tests per one million (the 7^th^ column of Table 1) as a criterion representing the level of facilities, therefore the quality of health care systems. Based on the information about this criterion, we define the lags (the distance between the test and diagnosis) for the different health care systems.

Table 2 represents the underlying properties of any country. As mentioned before, lag is the difference between the time of testing and the time of receiving the results of the tests, positive or negative, in days. The more facilities a health care system has, the more tests that system can do –therefore the lower positivity rate it has. Also, the more facilities a health care system has, the lower distance is between the tests and results. Based on the concept of lag, we pair the number of tests on the *n*^th^ day with the number of results on the (*n* + *lag*)^th^ day to obtain the dependency structure by using the copulas. The last column is calculated based on the start date and end date of the period of recording data (the fourth and fifth columns) and the lag (the sixth column), and it displays the number of pairs that we use to obtain the dependency structure for each country.

Generally, during an epidemic wave, the number of new infected individuals increases rapidly to an epidemic peak and then falls more gradually until the epidemic wave is over, and the number of new cases be stabilized. Roughly speaking, the epidemic peaks are the -neighborhood of-time points that corresponds the local maximum of the number of newly infected cases.

### Change point detection

We define the epidemic peak as the time neighborhood -or the time point-that *X*_*t*_:the number of new confirmed cases on the *t*^th^ day, exceeds the mean plus three times standard deviation of the last three weeks for at least a week, that is,

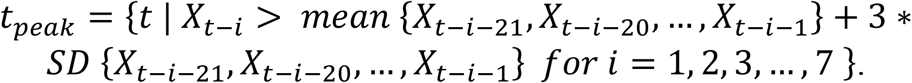

These epidemic peaks are local maximums. In addition, it is noticeable that the distance between two successive epidemic peaks must be at least one month. This definition is derived from the definition of outlier in regression analysis. According to this definition, the peaks of Table 3 are obtained for the countries of interest. It is remarkable that except for the peaks of Bolivia –which are almost the same-, the later waves are more acute than previous ones. We must add this point that the more acute peak means the more number of new confirmed cases, therefore the more intense spreading. Finally, it is possible that because of the lack of information at the beginning, this definition misses the epidemic peaks in the initial days.

**Table 3.**
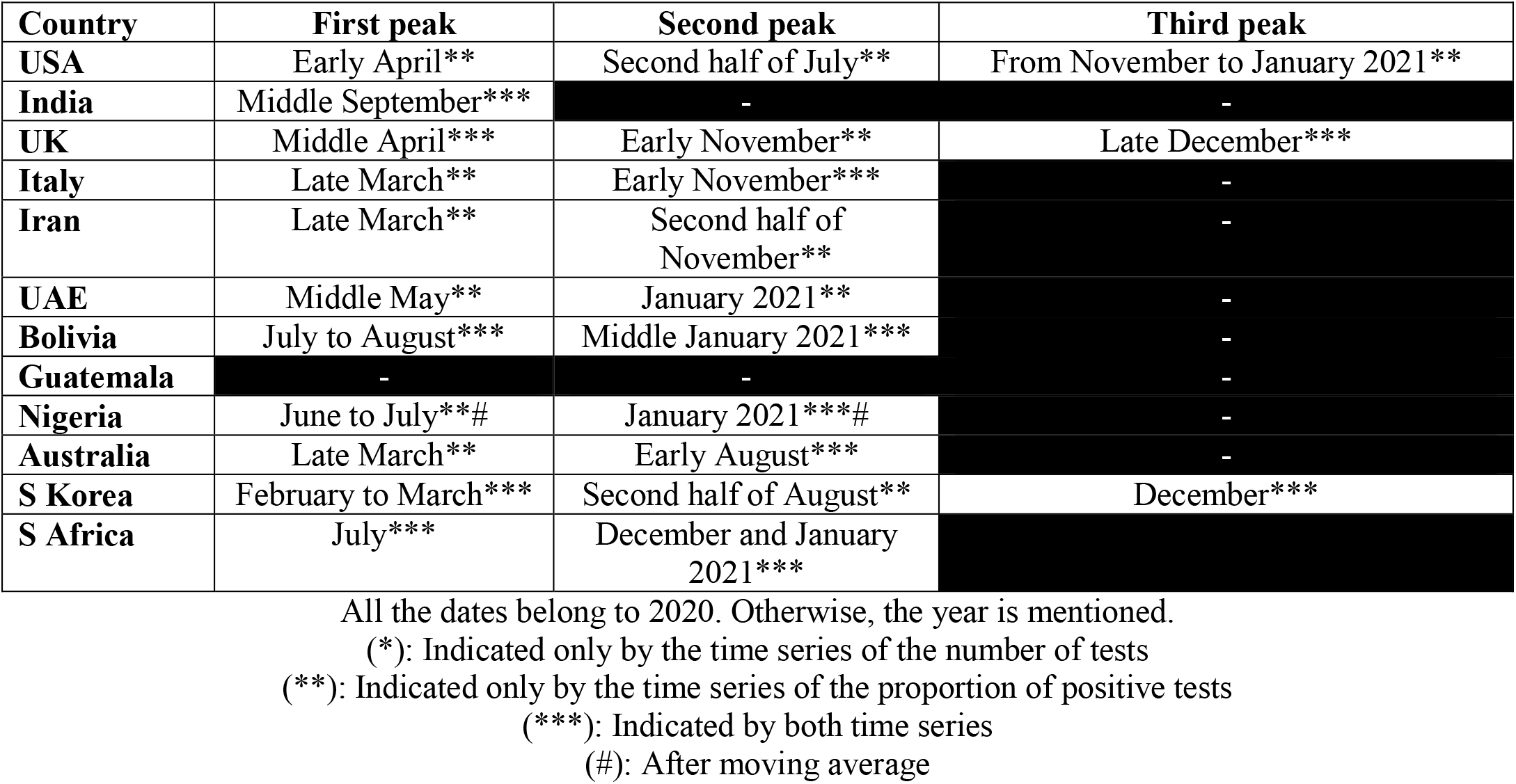
The epidemic peaks of COVID-19 in the countries of interest.

| Country | First peak | Second peak | Third peak |
| --- | --- | --- | --- |
| USA | Early April** | Second half of July** | From November to January 2021** |
| India | Middle September*** | - | - |
| UK | Middle April*** | Early November** | Late December*** |
| Italy | Late March** | Early November*** | - |
| Iran | Late March** | Second half of November** | - |
| UAE | Middle May** | January 2021** | - |
| Bolivia | July to August*** | Middle January 2021*** | - |
| Guatemala | - | - | - |
| Nigeria | June to July**# | January 2021***# | - |
| Australia | Late March** | Early August*** | - |
| S Korea | February to March*** | Second half of August** | December*** |
| S Africa | July*** | December and January 2021*** | - |

All the dates belong to 2020. Otherwise, the year is mentioned.
(\*): Indicated only by the time series of the number of tests
(\*\*): Indicated only by the time series of the proportion of positive tests
(\*\*\*): Indicated by both time series
(#): After moving average

**Table 4.** Kendall’s tau of copula function.

| Copula | Parameter space | Kendall's tau |
| --- | --- | --- |
| FGM | $\theta \in [-1, +1]$ | $\tau_k = 2\theta/9$ |
| Clayton | $\beta \in (0, +\infty)$ | $\tau_k = \beta/(\beta + 2)$ |
| Frank | $\alpha \in (-\infty, +\infty)$ | $\tau_k = 1 + 4D_{(\alpha)}/\alpha$, $D_{(\alpha)} = \frac{1}{\alpha} \int_0^\alpha \frac{x}{e^x - 1} dx - 1$ |
| Gumbel | $\sigma \in [1, +\infty)$ | $\tau_k = (\sigma - 1)/\sigma$ |

Mathematically and logically, the number of positive tests (confirmed cases) is affected by the number of tests and positivity rate. The number of cases equals the number of tests multiplied by the positivity rate. Therefore, the increment of the number of cases (as a multiplication) equals the sum of these two:

- The number of tests multiplied by the increment of positivity rate, and
- The positivity rate multiplied by the increment of the number of tests.

Consequently, the intense changes in the count of cases are due to at least a remarkable change in one of these multiplications. About the countries with a regular increase in the number of tests like the USA, the increment of the proportion of positivity plays the principal role in the peaks.

Table 3 shows that the proportion of positivity is significantly better than the frequency of tests to indicate the peaks of the pandemic. The positivity rate is more associated with the number of cases than the number of tests (90% versus 45%). After moving average, these proportions reach 100% and 50%, respectively.

Countries of the southern and northern hemispheres faced a peak around July and November, respectively, possibly due to falling temperatures.

Figures 1, 3, and 5 consist of twelve subfigures, each of them belonging to one country. The arrangement of the subfigures in all three figures is identical. The horizontal axes in Figures 1 and 3 represent time in days from the start to the end of the period of study for the studied countries (the fourth and fifth columns of Table 2). The vertical axis of Figures 1 and 3 display the number of new tests –conducted on that day-and the proportion of positive tests –reported that day-, respectively. Figure 5 is the plot of the joint distribution of the number of tests on a day and the proportion of positivity on the *lag* days later.

**Figure 1.**
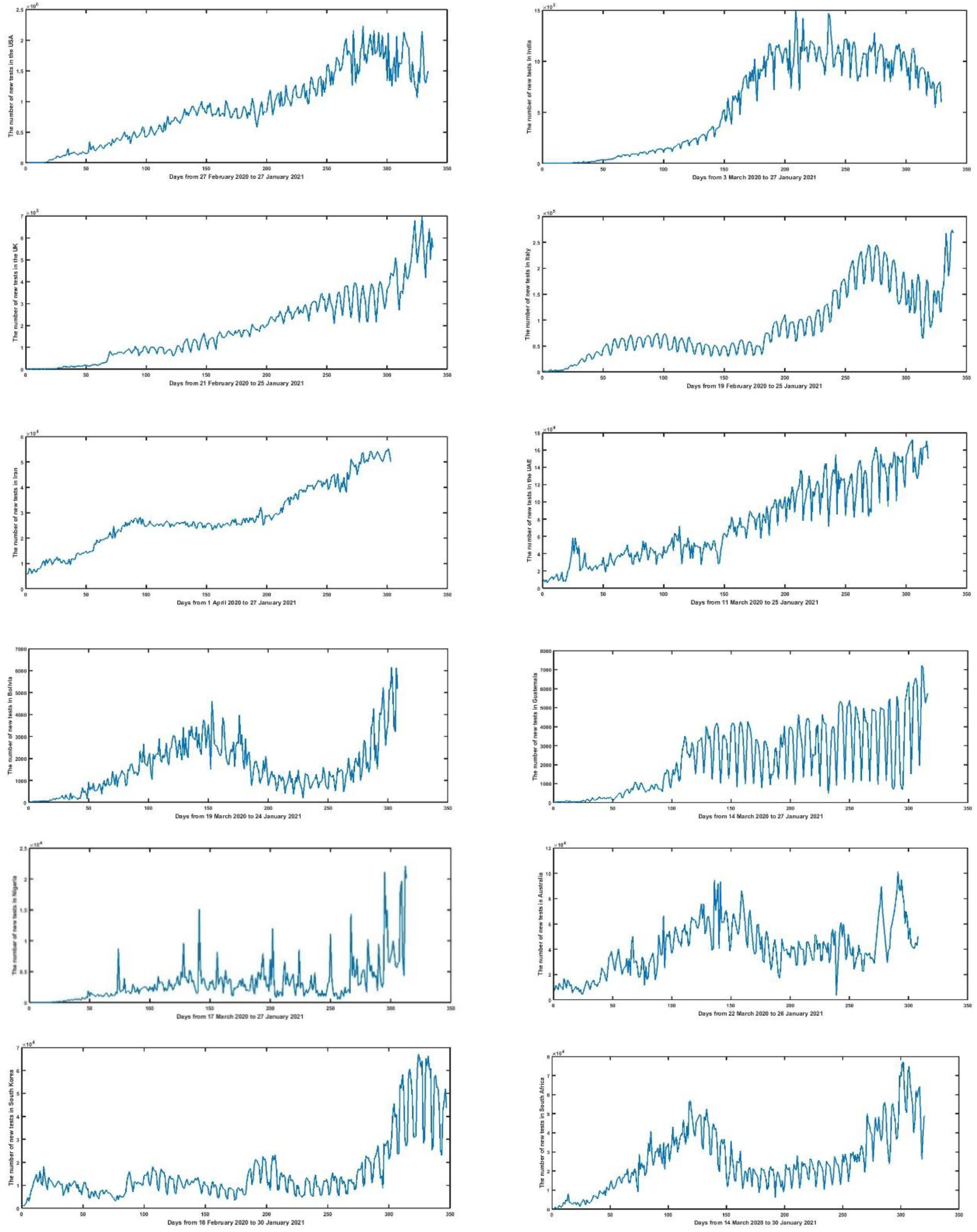
The time series of the number of new tests (daily) USA (r1 c1), India (r1 c2), UK (r2 c1), Italy (r2 c2), Iran (r3 c1), UAE (r3 c2), Bolivia (r4 c1), Guatemala (r4 c2), Nigeria (r5 c1), Australia (r5 c2), South Korea (r6 c1), and South Africa (r6 c2) r : row & c : column

**Figure 2.**
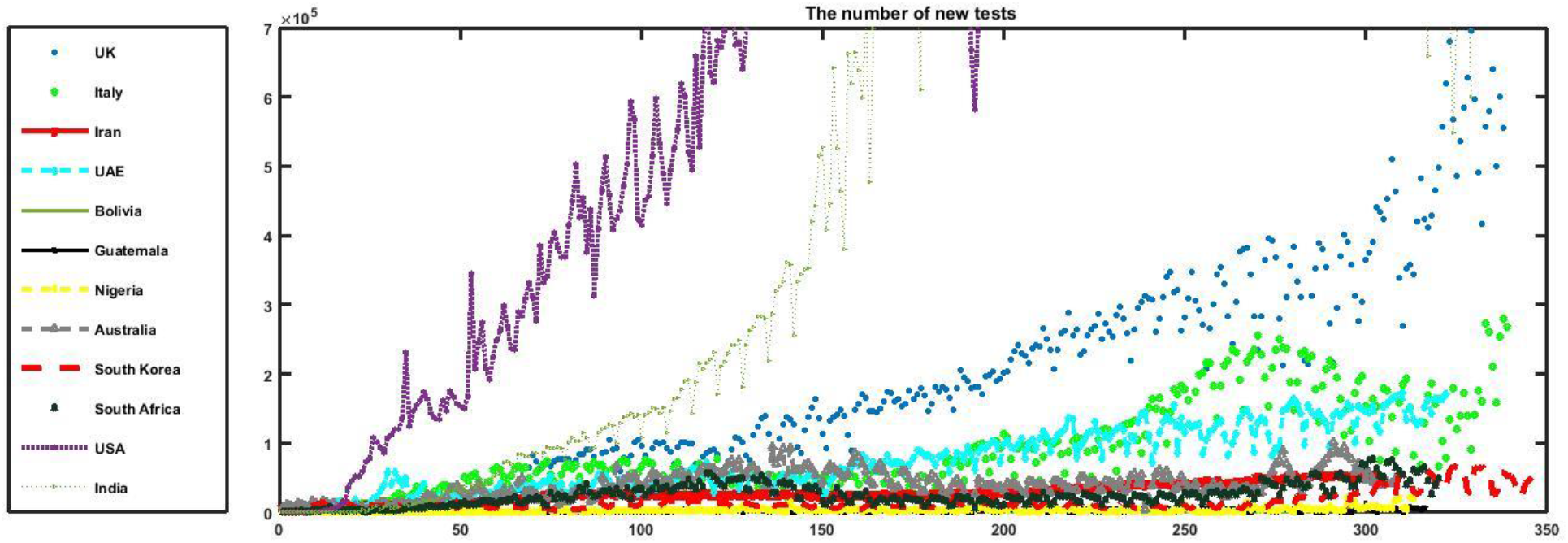
The time series of the number of new tests (daily)

**Figure 3.**
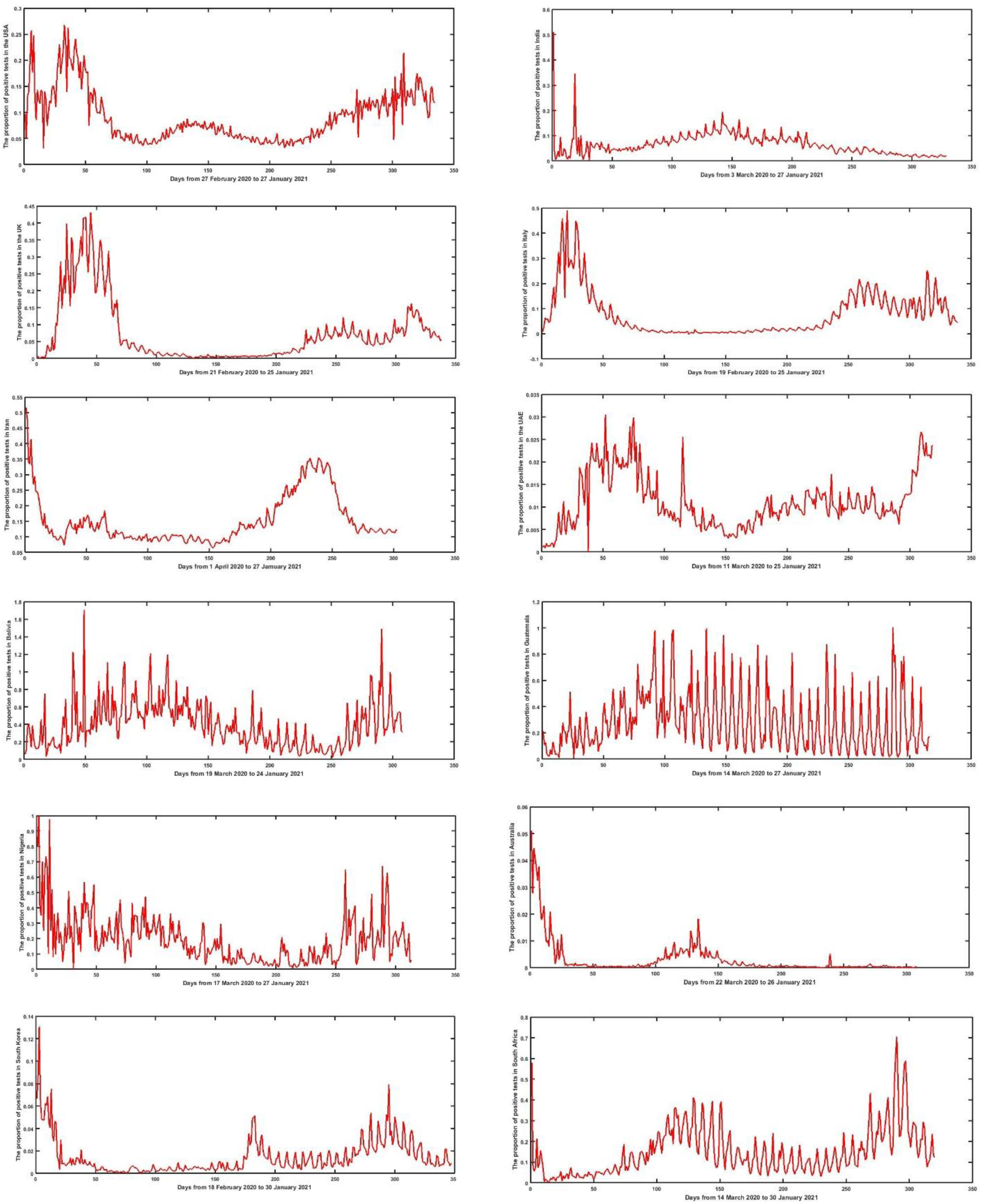
The time series of the positivity rate (daily) USA (r1 c1), India (r1 c2), UK (r2 c1), Italy (r2 c2), Iran (r3 c1), UAE (r3 c2), Bolivia (r4 c1), Guatemala (r4 c2), Nigeria (r5 c1), Australia (r5 c2), South Korea (r6 c1), and South Africa (r6 c2) r : row & c : column

**Figure 4.**
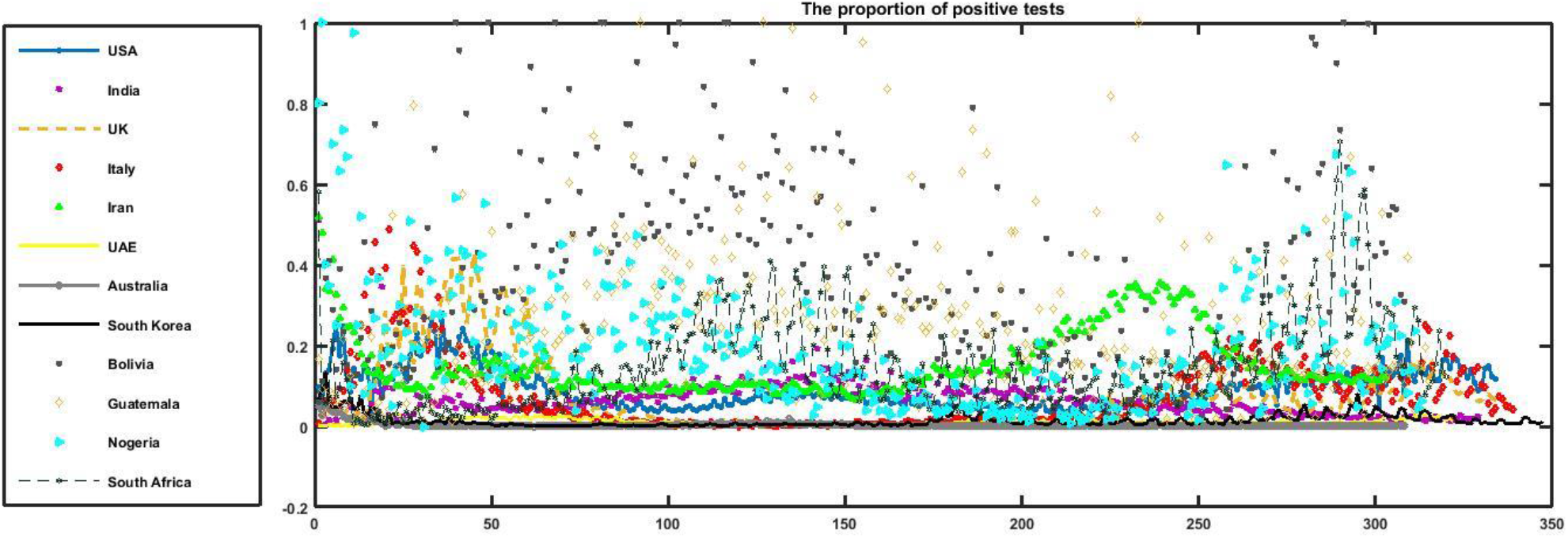
The time series of the positivity rate (daily)

**Figure 5.**
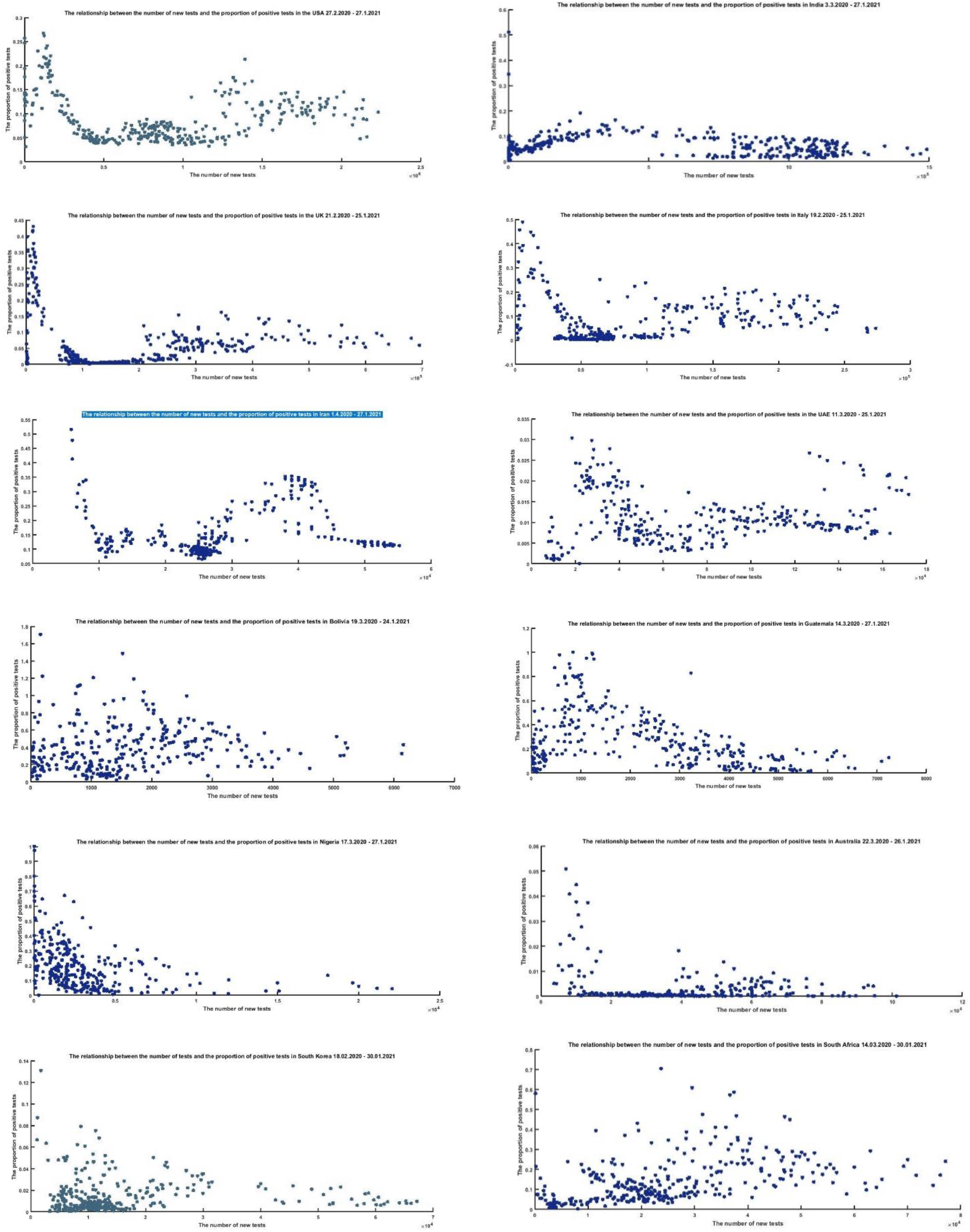
Scatterplots of the relationship between the number of tests and the positivity rate USA (r1 c1), India (r1 c2), UK (r2 c1), Italy (r2 c2), Iran (r3 c1), UAE (r3 c2), Bolivia (r4 c1), Guatemala (r4 c2), Nigeria (r5 c1), Australia (r5 c2), South Korea (r6 c1), and South Africa (r6 c2)

Figure 1 shows that the peaks of the number of tests coincide with the epidemic peaks of COVID-19 in different countries. For example, in the USA, there are two peaks of the number of tests simultaneous with the second and the third epidemic peaks –mentioned in Table 3-. Also, it is obvious that Bolivia has experienced two peaks for the number of tests around 150^th^ and 300^th^ days -from March 19, 2020-which coincide with the epidemic peaks in Table 3.

The USA, the UK, and the UAE experienced some regularly rising time series. Except for some overruns in epidemic peaks, the patterns of Italy and South Africa are increasing too. The number of tests in Guatemala is increasing, accompanied by an increasing fluctuation. Owing to the restriction by the limited capacity of tests, Iran and Nigeria followed a stepwise trend. Apart from the peaks, one for each of them, the plots of Australia and South Korea are stationary. In the case of Bolivia, the time series is proportional to the peaks. India is the only country whose time series is initially increasing, then stable, and after that decreasing. Generally, the counties have an increasing trend.

Figure 2 gives us a clustering about the countries from the viewpoint of the number of tests: 1. The USA, 2. India, 3. The UK, 4. Italy, Australia, and the UAE, 5. South Korea, South Africa, and Iran, and 6. Nigeria, Guatemala, and Bolivia.

Figure 3 illustrates the time series of the positivity rate of the tests (the ratio of the number of positive tests on a day to the number of taken tests on *lag* days ago). It is interesting that the subfigures of Figure 3 are more in accordance with the epidemic peaks than their analogous in Figure 1. For example, it is clear that the USA has encountered three peaks. It is worthwhile that the graph of Iran has three peaks while the first of them is missed in Table 3 because of the lack of information at the beginning. A similar situation (being missed by investigation of either the number of tests or the number of confirmed cases while discovered by the analysis of the positivity rate) happens to the epidemic peak in India in late March, the first and the second peaks of the UK, and the epidemic peaks in middle May and the November for the UAE.

Figure 4 illustrates a clustering of the countries based on the positivity rate: 1. Nigeria, Guatemala, and Bolivia, 2. South Africa, and Iran, 3. The USA, India, the UK, and Italy, and 4. Australia, the UAE, and South Korea.

The horizontal and vertical axes of Figure 5 display the number of new tests and the proportion of positivity of them, respectively. Generally, as the number of new tests increases, the positivity rate falls. Since the epidemic peaks are opposing this general rule, it is not very clear to see the opposite direction of the changes. Guatemala, due to lack of epidemic peak, is a good example of this diversely proportional relationship.

If the reason for an increase be the rising number of tests, we expect not to return the previous channel in short term. In addition, the positivity rate does not undertake a remarkable change. On the other hand, it is normal to assume that entering a peak is accompanied by increasing the number of negative tests as well. Consequently, the lack of the growth of negative test results (rising the positivity rate while continuing the previous trend for the frequency of tests) is only reasonable if at least one of the factors of tests accuracy, testing policy, or the viewpoint of the population were changed around that time. Otherwise, there are a remarkable number of un-reported cases belonging the peak. It is noticeable that this company of risings causes the observed acceleration in growth regarding epidemic peaks.

## Methods

### Copulas

Copulas are functions that connect multivariate distribution functions to their one-dimensional marginal distribution functions -uniform on the interval [0,1]. Mathematically speaking, if *H* is a bivariate distribution function with margins *F*(*X*) and *G*(*Y*), there must exist a copula *C* such that *H*_*θ*_ (*X, Y*) = *C*(*F*(*X*), *G*(*Y*);*θ*), where *θ* is introduced as the dependence parameter [5]. Accordingly, Copula is mostly defined as a function *C* : [0,1]^2^ → [0,1] that satisfies boundary conditions:

(P1) *C*(*x*,0) = *C*(0, *x*) = 0 and *C*(*x*,1) = *C*(1, *x*) = *x*, ∀*x* ∈[0,1],

(P2) ∀(*s*_1_, *s*_2_, *t*_1_, *t*_2_)∈[0,1]^4^, such that *s*_1_ ≤ *s*_2_ and *t*_1_ ≤ *t*_2_,

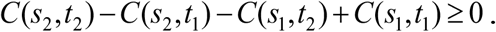

Eventually, for twice differentiable function *C*, 2-increasing property (P2) can be replaced by the condition

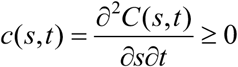

where *c*(*s,t*) is the so-called copula density. A copula *C* is *symmetric* if *C*(*s,t*) = *C*(*t, s*)

, for every(*s,t*) ∈[0,1]^2^, otherwise *C* is asymmetric. The most well-known, powerful, and applicable copulas are:

- FGM copula [19-20];

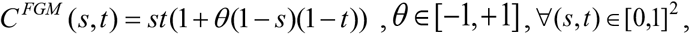

- Clayton copula [21];

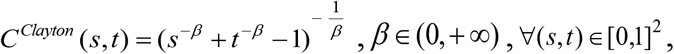

- Frank copula [22];

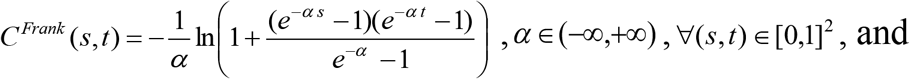

- Gumbel copula [23];

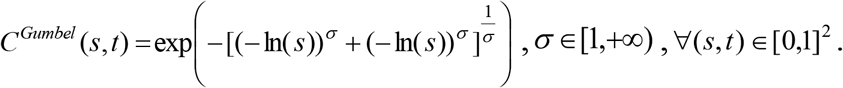

The parameters of the marginal and copula distributions are estimated using the maximum likelihood method. The computations and illustrations regarding copula theory are conducted in software Maple, R, and R 4.0.3, Maple 2018a, and EasyFit 5.6.

### Copula vs Correlation Coefficient

Measures of dependence are common instruments to summarize a complicated dependence structure in the bivariate case. Pearson’s, Spearman’s rho, and Kendall’s tau correlation coefficients are common statistical measures of dependence structure [24-26]. The correlation comes in trouble when the random variables are not elliptically distributed. The performance of the copula does not depend on the fact that if you are dealing with elliptical distributions or not. The Pearson’s linear correlation measure (−1≤ *r* ≤1) is the most popular and well-known measure between pairwise random variables. Despite its simplicity and plain rationale, Embrechts et al. [27] noted that *ρ* is simply a measure of the dependency of elliptical distributions, such as the binormal distribution (the marginals are normally distributed, linked by the Gaussian copula). Moreover, *ρ* measures a linear relationship itself and does not capture a non-linear one on its own, as noted in [28]. These properties constitute obvious limitations for modeling the dependency structure. In addition, copulas could be useful to define nonparametric measures of dependence between random variables. Since the values of Kendall’s tau are easy to calculate, this measure is used for observation dependencies. If *F*(*X*) and *G*(*Y*) are continuous then *C*(*s,t*) is unique, else *C*(*s,t*) is uniquely determined on the range of *F*(*X*) × range of *G*(*Y*).

One standard non-parametric dependence measures Kendall’s *τ* _*k*_ is expressed in the copula form as:

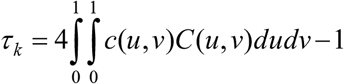

The parameter copula is estimated and the relationship between parameter copula and *τ* _*k*_ is given in the last column of Table 1. The parameter copula in each case measures the degree of dependence and controls the association between two variables. When the parameter approaches 0 there is no dependence, and if the parameter tends to infinity there is a perfect dependence. Schweizer and Wolff [29] showed that the dependence parameter copula, which characterizes each family of copulas can be related to Kendall’s *τ*_*k*_. Therefore, copulas allow modeling both linear and non-linear dependence. Using copulas, regardless of marginal distributions, can model extreme endpoints.

### Copula vs Regression

Regression analysis is a statistical method for investigating the relationships between some dependent variables and some independent variables. The basic form of the regression analysis, ordinary least squares is not suitable for some applications because the relationships are often nonlinear and the probability distribution of the response variable may be non-Gaussian.

The major advantage of copula regression is that there are no restrictions on the probability distributions that can be used. The copula regression is the most appropriate method in non-Gaussian (no need for normality assumption) regression model fitting. Copula functions, connecting the marginal distributions to their joint distributions, are useful in simulating the linear or nonlinear relationships among multivariate data. Copula is a multivariate distribution function with marginally uniform random variables on [0, 1] (the PDF of the CDF). Copula functions have some appealing properties such as they allow scale-free measures of dependence and are useful in constructing families of joint distributions.

## Results

The presumptions to apply copula theory for a couple of variables are the existence of continuous marginal distributions accompanied with their correlation. Table 5 investigates whether the pair of the frequency of the tests and positivity rate meets the presumptions. The marginal distributions were obtained in EasyFit. It is observable that the generalized Pareto and Weibull distributions had good performance to fit the positivity rates. It is observable that the correlation in countries with the highest number of tests is negative and it is commonly between −0.2 and - In countries lacking enough tests, the correlation coefficient is significantly greater –possibly due to the low quality of data and under-reporting. It is noticeable that calculation over the data of Bolivia, Iran, and South Africa, lead even to positive correlations.

**Table 5.** The results of fit distribution to data.

| Country | frequency of tests data |  |  |  | positivity proportion data |  |  |  | Correlation |  |
| --- | --- | --- | --- | --- | --- | --- | --- | --- | --- | --- |
| | Marginal | Parameters | K-S Test | | Marginal | Parameters | K-S Test | | $r$ | P-Value |
|  |  |  | Statistic | P-Value |  |  | Statistic | P-Value |  |  |
| **USA** | Rayleigh | $\sigma = 864763$<br>$\gamma = -195972$ | 0.0654 | 0.11266 | Gen. Pareto | $k = -0.14127$<br>$\sigma = 0.06493$<br>$\mu = 0.03461$ | 0.04538 | 0.48317 | -0.134 | 0.014 |
| **India** | Logistic | $\sigma = 252223$<br>$\mu = 57602$ | 0.04919 | 0.19214 | Weibull | $\alpha = 1.8788$<br>$\beta = 0.07082$ | 0.04234 | 0.58217 | -0.236 | < 0.01 |
| **UK** | Gen. Pareto | $k = -0.37332$<br>$\sigma = 280831$<br>$\mu = -14012$ | 0.05684 | 0.2165 | Weibull (3p) | $\alpha = 0.78036$<br>$\beta = 0.06417$<br>$\gamma = 0.00833$ | 0.07206 | 0.05687 | -0.213 | < 0.01 |
| **Italy** | Log-Logistic (3P) | $\alpha = 2.6282$<br>$\beta = 87799.0$<br>$\gamma = -16892.0$ | 0.06078 | 0.15679 | Weibull (3p) | $\alpha = 0.86458$<br>$\beta = 0.07299$<br>$\gamma = -0.00238$ | 0.07386 | 0.0521 | -0.001 | 0.986 |
| **Iran** | Log-Logistic (3P) | $\alpha = 8.1298$<br>$\beta = 56499.0$<br>$\gamma = -29429.0$ | 0.00736 | 0.05101 | Burr | $k = 0.16689$<br>$\alpha = 13.051$<br>$\beta = 0.08904$ | 0.05516 | 0.3040 | 0.123 | 0.032 |
| **UAE** | Weibull | $\alpha = 1.5811$<br>$\beta = 84164.0$ | 0.05992 | 0.19016 | Log-Logistic | $\alpha = 3.0628$<br>$\beta = 0.01041$ | 0.07394 | 0.05619 | -0.001 | 0.854 |
| **Bolivia** | Gumbel Max | $\sigma = 923.72$<br>$\mu = 1072.6$ | 0.04872 | 0.44386 | Beta | $\alpha_1 = 1.3627$<br>$\alpha_2 = 2.9923$ | 0.0332 | 0.87483 | 0.189 | 0.001 |
| **Guatemala** | Dagum | $k = 0.0587$<br>$\alpha = 10.772$<br>$\beta = 5929.2$ | 0.0707 | 0.08088 | Gamma | $\alpha = 1.9352$<br>$\beta = 0.13129$ | 0.03456 | 0.8318 | -0.329 | < 0.01 |
| **Nigeria** | Log-Logistic | $\alpha = 2.3097$<br>$\beta = 2736.4$<br>$\gamma = -510.36$ | 0.04696 | 0.48053 | Weibull | $\alpha = 1.2881$<br>$\beta = 0.2093$ | 0.03527 | 0.81772 | -0.371 | < 0.01 |
| **Australia** | Logistic | $\sigma = 11052.0$<br>$\mu = 40909.0$ | 0.04808 | 0.46066 | Frechet | $\beta = 0.0043$<br>$\alpha = 0.77645$ | 0.05106 | 0.38529 | -0.269 | < 0.01 |
| **S Korea** | Burr | $k = 0.53449$<br>$\alpha = 3.5823$<br>$\beta = 8601.9$ | 0.05161 | 0.30334 | Gen. Pareto | $k = 0.18396$<br>$\sigma = 0.01109$<br>$\mu = 0.00924$ | 0.03947 | 0.63718 | -0.005 | 0.926 |
| **S Africa** | Log-Logistic (3P) | $\alpha = 4.5531$<br>$\beta = 40005.0$<br>$\gamma = -17075.0$ | 0.03938 | 0.68861 | Gen. Pareto | $k = -0.13432$<br>$\sigma = 0.14687$<br>$\mu = 0.01868$ | 0.02748 | 0.96362 | 0.405 | < 0.01 |

K-S: Kolmogrov-Smirnov.
3p: 3-parameter.
The highlighted rows indicate that the correlation are not significant for those countries.

Based on Table 5, we are allowed to look for the suitable copula functions to connect the marginal distributions to find the desired joint distributions for nine of the countries. Notice that the countries without meaningful correlation (Italy, South Korea, and the UAE) were of the countries with the least proportion of positivity of the tests. These countries have involved with tracing the infected cases.

Table 6 represents the results of comparing the best candidates from the FGM, Clayton, Frank, and Gumbel families.

**Table 6.**
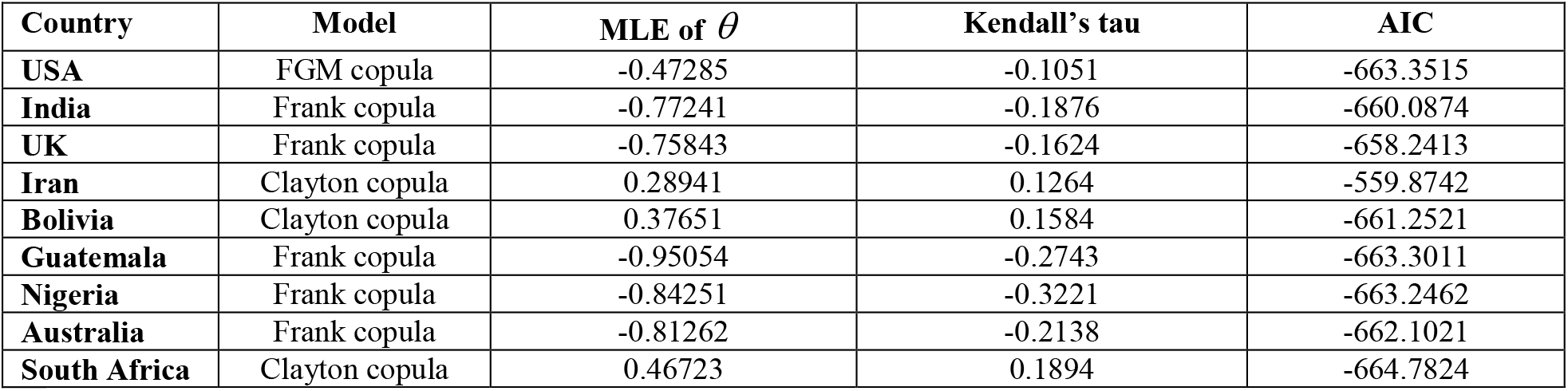
The obtained copula to fit the dependency and their performances.

According to Table 6, Clayton copulas are suitable candidates for the countries with low tests per million. In addition, Frank copulas can describe a wide variety of countries. Finally, the Gumbel family seems not to be a good option to couple the variables of the frequency of tests and the positivity rate.

We now discuss the simulation of data from the obtained copula models and perform comparisons between correlations in the simulated data and in the observed data based on 1000 simulations. We follow the simulation method proposed by Johnson (1987, Ch.3) and later Nelson (2006, page 41).

Figure 6 illustrates the scatter plots of the transformed observed data versus simulated samples of the CDFs of the frequency of tests and positivity proportion variables taken from the fitted copula models in Table 6. It can be seen that the simulated data and the original data have similar dependence patterns. To settle this concern, Table 6 shows the rank correlations between the frequency of tests and positivity proportion variables calculated from the original data and the simulated data of size 1000 taken from the fitted copula models. By comparing these correlations, we can conclude that the results show strong consistency of the estimated and real correlations.

**Figure 6.**
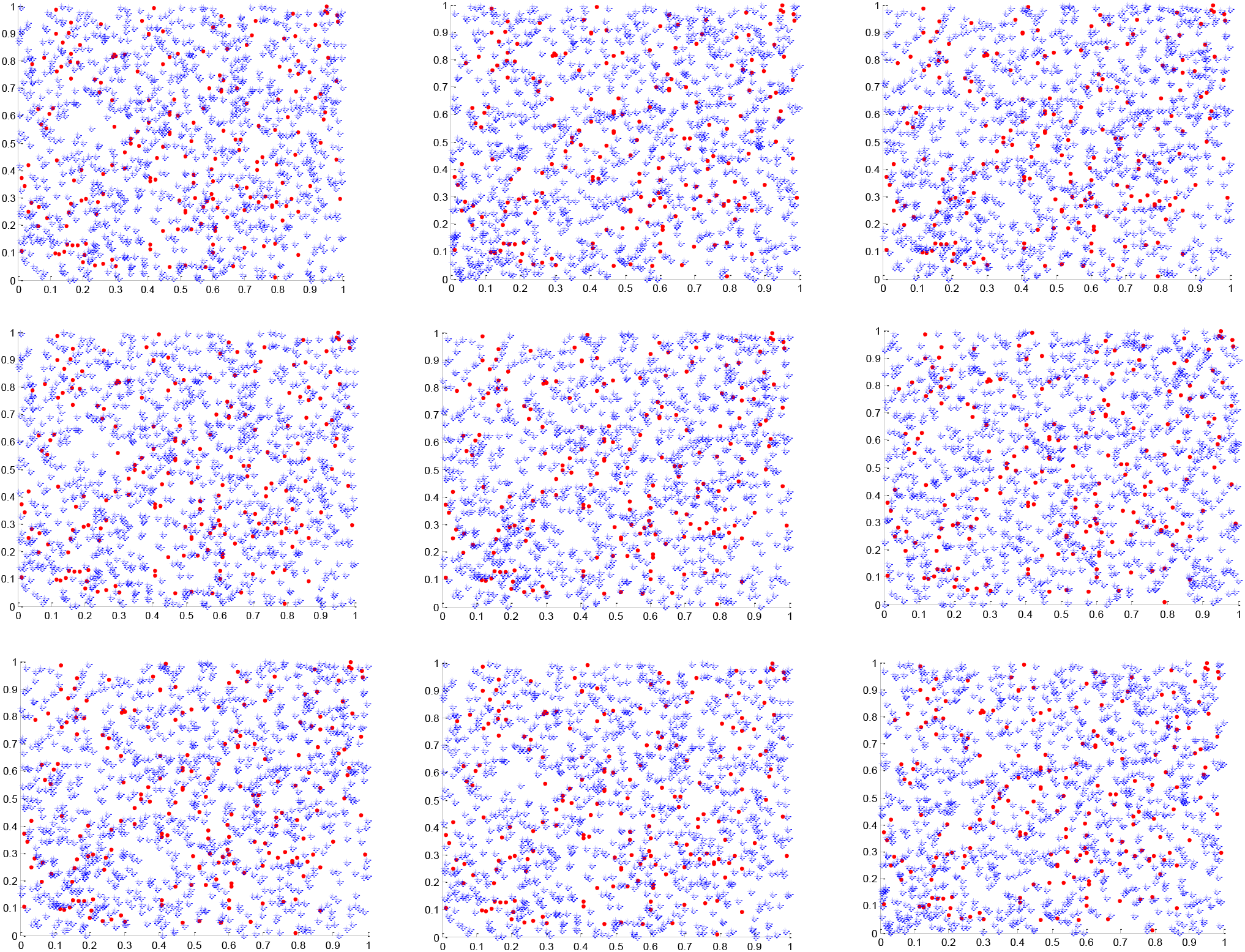
Scatter plots of the transformed observed values (•) versus simulated samples (∗) variables from subfamilies of the copula model USA (r1 c1), India (r1 c2), UK (r1 c3), Iran (r2 c1), Bolivia (r2 c2), Guatemala (r2 c3), Nigeria (r3 c1), Australia (r3 c2), and South Africa (r3 c3) r : row & c : column

Finally, we want to investigate the structure of dependency between the number of tests and positivity rate totally. By collecting the data of the twelve countries, 3877 pairs are obtained whose Kendall’s correlation is −0.1434 (P-value: 2.8464*10^ −19). In addition, we split the data into two parts: peaks and otherwise. This split restricted us to applying marginal distributions –then copulas_ because it causes the gap in the number of tests. Table 7 represents the Kendall’s correlations for the countries of interest. It is worth saying that the correlation coefficient for the variables (the number of tests and positivity rate) is negative in both peaks and otherwise.

**Table 7.** The correlation between the number of tests and the positivity rate regarding all countries separated based on the peaks.

| Country | Kendall's tau after removing peaks | P-value | Kendall's tau for peaks | P-value |
| --- | --- | --- | --- | --- |
| USA | -0.0168 | 0.8113 | **-0.4104** | **1.2402E-6** |
| India | **-0.2410** | **1.0457E-4** | 0.0993 | 0.3936 |
| UK | 0.2496 | 0.0015 | **-0.7017** | **1.2403E-25** |
| Italy | 0.3309 | 2.0731E-7 | **-0.5127** | **5.8909E-11** |
| Iran | 0.0779 | 0.2354 | 0.1574 | 0.1898 |
| UAE | 0.0387 | 0.5348 | -0.1621 | 0.2081 |
| Bolivia | 0.3028 | 8.7577E-6 | **-0.2402** | **0.0119** |
| Guatemala | **-0.2946** | **9.5948E-8** |  |  |
| Nigeria | 0.4474 | 2.3797E-9 | **-0.4197** | **6.6622E-8** |
| Australia | **-0.2337** | **3.1165E-4** | **-0.6295** | **3.1617E-9** |
| South Korea | 0.3134 | 8.5238E-8 | **-0.7214** | **5.7158E-12** |
| South Africa | 0.1203 | 0.0744 | **-0.4517** | **22.3897E-6** |
| Total | **-0.1381** | **3.0125E-13** | **-0.2132** | **2.9617E-13** |

Light or dark bolded figures indicate that the coefficient correlation is significantly positive or negative, respectively.

## Discussion

Generally, at the beginning of an epidemic, the number of tests is low and the proportion of positivity is high. As time passes, the number of tests rises. Also, as the number of new tests increases, the positivity rate falls. The correlation in countries with high number of tests, higher quality of data, is negative and it is commonly between −0.2 and −0.3. By considering all the data as a set, the Kendall’s coefficients are −0.1434, −0.2132, and −0.1381 for total, peaks, and total after removing peaks, respectively. The positivity rate of the tests is significantly better than the frequency of tests to indicate the peaks of the pandemic. The positivity rate is more associated with the number of cases than the number of tests (90% versus 45%).

The proportion of positivity is more proportional than the number of tests to the number of infected cases. Approaching zero positivity rate is a good criterion to scale the success of a health care system in fighting against an epidemic. The number and accuracy of tests can play a vital role in the quality level of epidemic data. The policymakers can consider the factors affecting the positivity rate such as the testing policy, restricted facilities, peaks, fluctuations, and so on, and make decisions to prevent misleading because of them.

The first limitation is the low quality of data for some countries because of the restricted facilities, the low number of tests, and non-organized data collection program. Also, some interpolation and moving average methods were applied to find some missing data regarding the countries of interest and calculating the correlation for the countries with poor data. Out of the twelve countries, Iran, South Africa, Nigeria, Bolivia, and Guatemala have been restricted by the number of tests. The data of Italy, the UAE, and South Korea showed no significant correlation. The lack of dependency is a good criterion to show that there is no shortage of facilities. The highest quality and most significant correlations belong to the USA, India, the UK, and Australia.

The present approach using copulas is promising since it allows to take into account a wide range of correlation, frequently observed in medical. In fact, the classical multivariate models cannot reproduce all type of correlations. Moreover, the standard models are limited, especially because the choice of the marginal distributions is restricted. The crucial step in the modeling process is the choice of the copula function, which best fits the data. Further work is needed to choose the best copulas able to reproduce the dependence structure of bivariate medical variables. In clinical trials or medical studies, sample size is often an important consideration and is relatively small. The copula-based methodology overcomes this limitation as well, because the algorithm can be used to replicate data for any number of patients. The suggested copula-based methodology presented in this paper is simple and easy to implement.

## Data Availability

The main data can be found on https://www.worldometers.info/coronavirus/countries and
https://ourworldindata.org/coronavirus-testing

## Ethics declarations

### Ethical statement

The methodology for this study was approved by the Human Research Ethics committee of the Kermanshah University of Medical Sciences.

### Informed consent

It is not applicable. This study did not deal with human and animal subjects.

### Consent on publication

This is not applicable. The manuscript includes no case study.

## Acknowledgment

We are grateful to Azad Sheikhi for helping us to write better in English.

## Contributorship

**BJ:** Idea, Literature, Data, Methods, Programming, Interpretation, First draft. **HB:** Literature, Methods, Interpretation, Revision. **SJZ:** Data, Literature, Programming. **MR:** Design, Final manuscript.

## Data availability

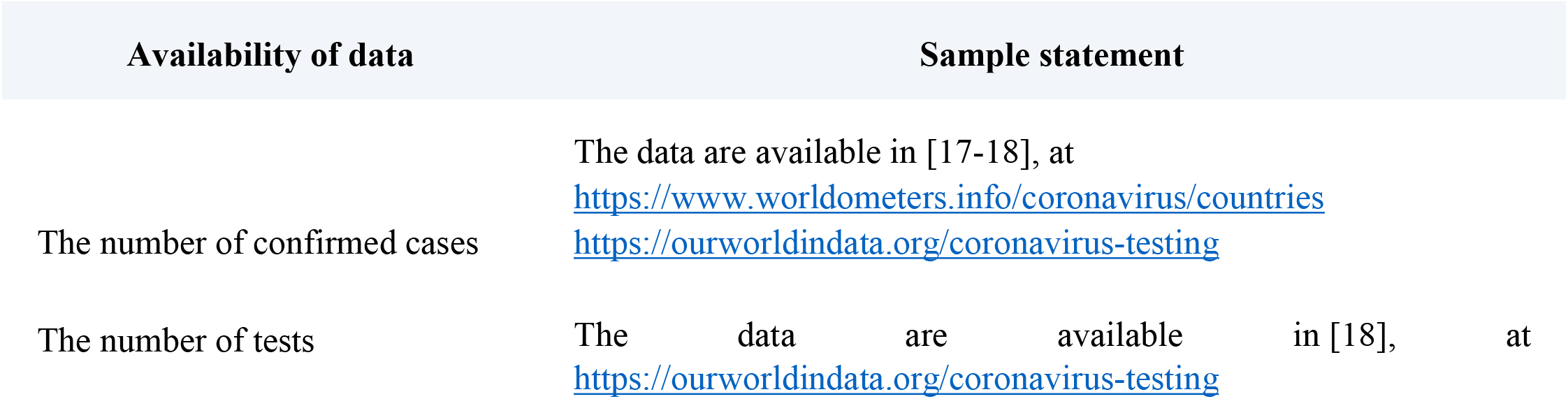

| Availability of data | Sample statement |
| --- | --- |
| The number of confirmed cases | The data are available in [17-18], at<br>https://www.worldometers.info/coronavirus/countries<br>https://ourworldindata.org/coronavirus-testing |
| The number of tests | The data are available in [18], at<br>https://ourworldindata.org/coronavirus-testing |

